# Health-related effects of education level: a Mendelian randomization study

**DOI:** 10.1101/2020.02.01.20020008

**Authors:** Shuai Yuan, Ying Xiong, Madeleine Michaëlsson, Karl Michaëlsson, Susanna C. Larsson

## Abstract

**Background:** A deeper understanding of the causal links from education level to health outcomes may shed a light for disease prevention at a novel and efficient perspective.

**Methods:** We conducted a wide-angled Mendelian randomization to disentangle the causal role of education level from intelligence for 31 health outcomes and explore to what extent body mass index and smoking mediate the associations. Univariable and multivariable Mendelian randomization analyses were performed.

**Results:** Genetically higher education level was associated with lower risk of major mental disorders and most somatic diseases independent of intelligence. The intelligence-adjusted odds ratios for each additional standard deviation of education (4.2 years) were 0.48 (0.37, 0.62) for suicide attempts, 0.50 (0.36, 0.68) for large artery stroke, 0.51 (0.42, 0.63) for heart failure, 0.52 (0.42, 0.65) for lung cancer, 0.45 (0.33,0.61) for rheumatoid arthritis, and 0.48 (0.43, 0.55) for type 2 diabetes. Higher education level adjusted for intelligence was additionally associated with lower risk of insomnia, major depressive disorder, stroke, coronary artery disease, breast cancer, ovarian cancer and gout but with higher risk of obsessive-compulsive disorder, anorexia nervosa, bipolar disorder and prostate cancer. Moreover, higher education level was associated with modifiable health-related risk factors in a favorable manner. Adjustment for body mass index and smoking attenuated the associations between education level and several outcomes, especially for type 2 diabetes and heart failure. High education level exerts causal protective effects on major somatic diseases.

**Conclusions:** These findings emphasize the importance of education to reduce the burden of common diseases.

## Main text

Education level is an important health social determinant and has been proposed as a modifiable risk factor for a number of disorders and diseases, such as depression ^1^, age-related cognitive decline ^2^, suicide ^3^, cardiovascular disease ^4^, cancer ^5^, and several other diseases ^6-8^. However, it is unclear whether the associations are causal and independent of intelligence. Understanding the causal effects of education level on diseases can facilitate the aetiology pathway exploration of diseases as well as development of new strategies for disease prevention. Notwithstanding, randomized controlled trials are ethically and practically infeasible on this topic.

Exploiting genetic variants as instrumental variables for an exposure (e.g., education level), Mendelian randomization (MR) can strengthen the causal inference of an exposure-outcome association ^9^. Comparing the risk of disease across individuals who have been classified by their genotype enables the causal effect of an exposure to be estimated with substantially less bias, such as confounding and reverse causality, than in a traditional observational analysis ^9^. The rationale for diminished bias in MR studies is that genetic variants are randomly assorted and fixed at conception and therefore largely independent of confounders and cannot be modified by disease development ^9^.

We conducted an MR study to disentangle the causal role of education level from intelligence in major mental and neurological disorders and somatic diseases. A secondary aim was to explore whether intelligence is causally associated with the same health outcomes independently of education. We additionally investigated the associations of education level and intelligence with modifiable health-related risk factors and whether main lifestyle factors mediate the pathway from education to health outcomes.

## Methods

### Study design

The design and hypothesis of the present study are displayed in Supplementary Figure 1. We used summary-level data from large genome-wide association studies (GWASs) and genetic consortia (Table 1). Totally, our study included 11 mental and neurological disorders ^10-20^, 20 major somatic diseases ^21-36^, and 10 health-related risk factors ^31,34,35,37-39^. A systematic review was conducted to find meta-analyses of observational studies of education level and diseases (Supplementary table 1).

**Table 1.**
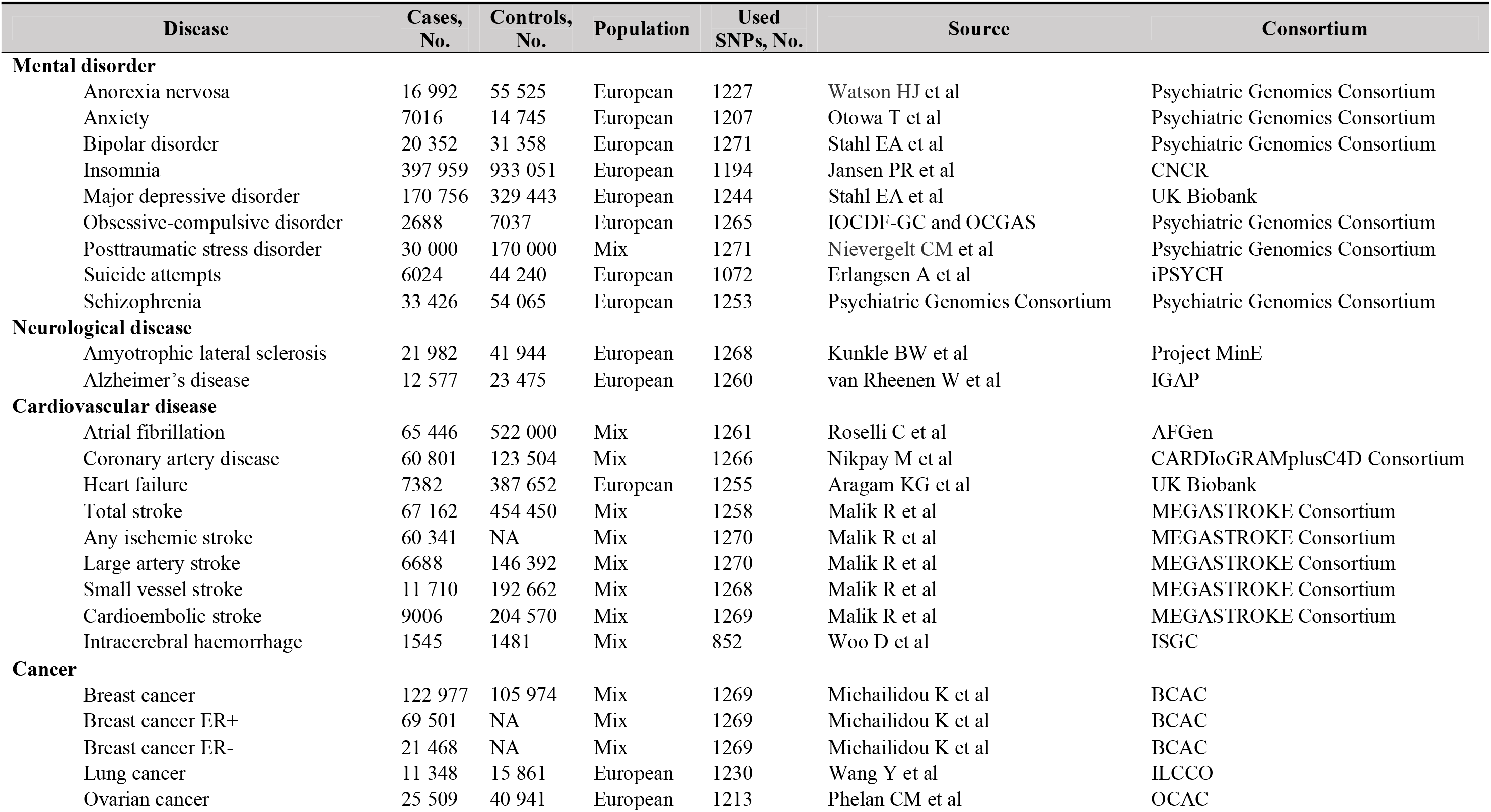
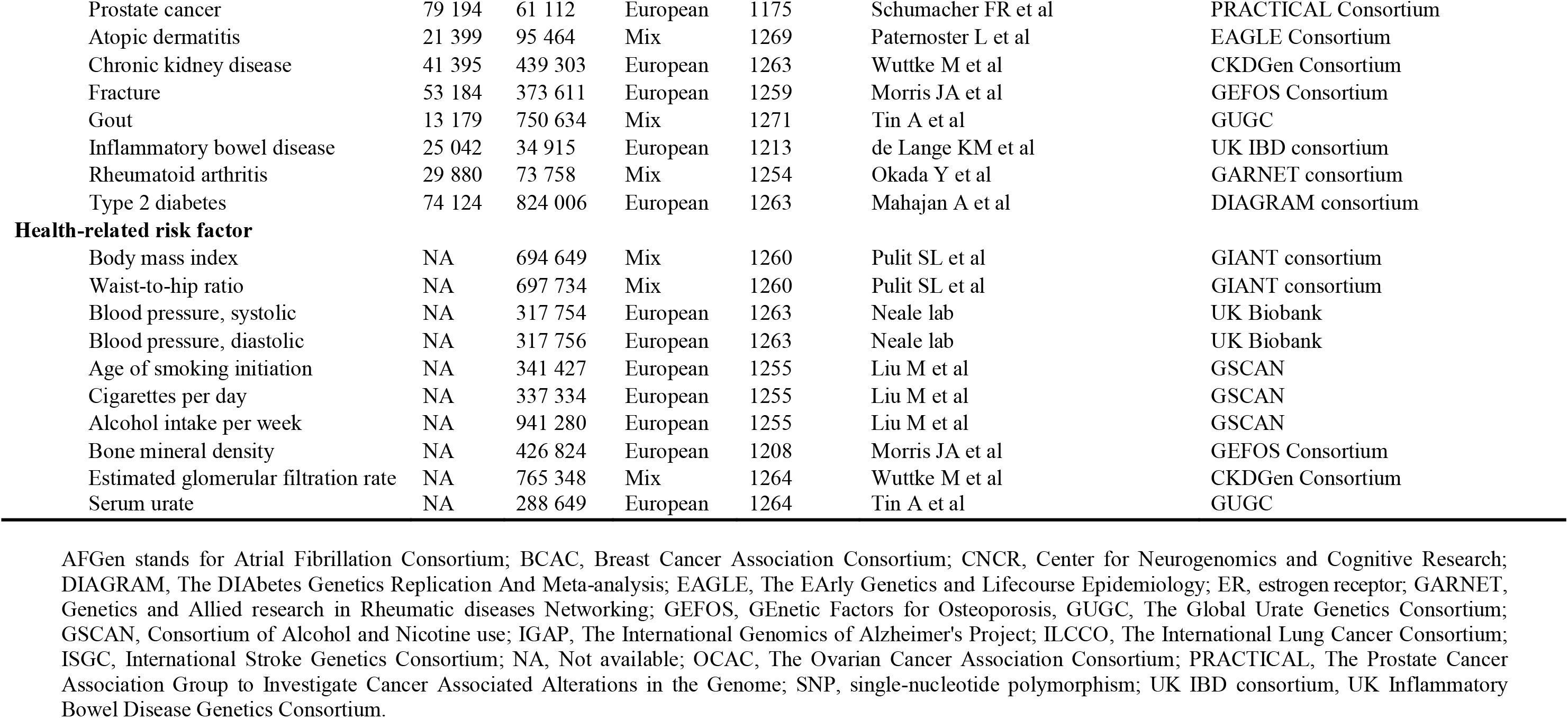
Characteristics of included studies of mental disorders, somatic diseases, and health-related risk factors

### Selection of instrumental variables

Instrumental variables for education level and intelligence were identified from GWASs of, respectively, 1 131 881 and 269 867 individuals of European ancestries ^40,41^. In total, 1271 and 205 single-nucleotide polymorphisms (SNPs) at the genome-wide significance threshold (*p*<5×10^-8^) were proposed as instrumental variables for education level and intelligence. All used SNPs explained around 12.2% and 5.2% variance for education level and intelligence, respectively. Education level was defined as number of years of education and was unified across included studies according to an International Standard Classification of Education category. Across all cohorts, the sample-size-weighted mean of education year was 16.8 years of schooling with a standard deviation (SD) of 4.2 years. For the definition of intelligence as an instrumental variable in our analysis, included cohorts extracted a single sum score, mean score, or factor score from a multidimensional set of cognitive performance tests in GWAS with linear model, with the exception of High-IQ/Health and Retirement Study where a logistic regression GWAS was run with “case” status (high intelligence) versus controls (normal intelligence level). All included GWASs adjusted for key covariates, such as age, sex and principal components for ancestry. The numbers of SNPs used as instrumental variables in each analysis are displayed in Table 1.

### Outcome sources

Summary-level data for the associations of the education- and intelligence-associated SNPs with the outcomes were extracted from large-scale GWASs or genetic consortia. In the present MR study, we included 11 mental and neurological disorders ^10-20^, 9 cardiovascular diseases ^21-25^, 4 major cancers ^26-29^, 7 other diseases ^30-36^ and 10 established health-related risk factors for major diseases ^31,34,35,37-39^. Detailed information, such as the number cases and controls, population structure and the source for each outcome, is presented in Table 1. Definitions of the diseases are presented in Supplementary Table 2.

### Systematic review for meta-analysis of observational studies

A systematic literature search was conducted in the PubMed database before November 1^st^, 2019 to find meta-analyses of observational studies of education level in relation to diseases studied in the present MR study. We found latest published meta-analysis on 13 diseases and two risk factors, including major depressive disorders ^42^, suicide attempts ^43^, posttraumatic stress disorder ^44^, amyotrophic lateral sclerosis ^45^, Alzheimer’s disease ^46^, coronary artery disease ^47^, heart failure ^48^, stroke ^49^, breast cancer ^50^, prostate cancer ^51^, lung cancer ^52^, type 2 diabetes ^53^, chronic kidney disease ^54^, body mass index ^55^ and hypertension (blood pressure) ^56^. We extracted publication data (PMID number, the first author’s name and year of publication), sample size, and risk estimates with their corresponding confidence intervals. Search strategy and characteristics of included meta-analyses are shown in Supplementary Table 1.

### Statistical analysis

The random-effects inverse-variance weighted method was used to assess the associations of education and intelligence with the outcomes. The weighted median method and MR-Egger regression were used as sensitivity analyses to examine the consistency of results and to detect potential pleiotropy. The weighted median method gives accurate estimates if at least 50% of the instrumental variables are valid ^57^. The MR-Egger regression can detect and adjust for pleiotropy albeit rendering low precision of the estimates ^58^. Given the phenotypical and genetic correlation between education level and intelligence ^59^, we used a multivariable inverse-variance weighted method to disentangle the causal effect of education level on outcomes independent of intelligence and vice versa ^60^. Considering that smoking and body mass index are modifiable risk factors linking education level to the most diseases, we also used multivariable-adjusted MR analysis to determine the effects of body mass index and smoking behavior on health outcomes for associations reaching the conventional significance level (*p*<0.05) in both univariable and intelligence-adjusted inverse-variance weighted model to explore the mediation effects of body mass index and smoking ^61^. Proportions of attenuated effect size were calculated to present the magnitude of mediation effects. Odds ratios (ORs) and 95% confidence intervals (CIs) of diseases and changes of levels of risk factors were scaled to an SD increase in genetically predicted years of education (4.2 years) and intelligence. We calculated the power for the analyses of education level using a web-tool ^62^ and results are displayed in Table 1. All statistical analyses were two-sided and performed using the mrrobust package in Stata/SE 15.0 ^63^ and TwoSampleMR in R 3.6.0 software and MR-Base ^64^. *P* values were not used strictly to define statistical significance; however, we interpreted the results based on the magnitude and strengths of the associations ^65^.

## Results

### Genetically predicted education level and diseases

Genetically predicted education level was causally associated with most diseases, including 8 out of 11 mental and neurological disorders, all 9 studied cardiovascular diseases, all 4 studied cancers, and 4 out of 7 other common diseases in the univariable inverse-variance weighted MR analysis (Figure 1). In multivariable inverse-variance weighted analysis with adjustment for intelligence, higher education level was additionally associated with higher odds of schizophrenia and anxiety (these associations were not observed in the univariable analysis) (Figure 1). However, the inverse associations of education level with amyotrophic lateral sclerosis, Alzheimer’s disease, cardioembolic stroke, intracerebral haemorrhage and inflammatory bowel disease observed in the crude MR analysis did not remain after adjustment for intelligence (Figure 1). In the intelligence-adjusted model, higher education level was associated with lower odds of rheumatoid arthritis (OR 0.45; 0.33, 0.61), type 2 diabetes (OR 0.48; 0.43, 0.55), suicide attempts (OR 0.48; 0.37, 0.62), large artery stroke (OR 0.50; 0.36, 0.68), heart failure (OR 0.51; 0.42, 0.63), lung cancer (OR 0.52; 0.42, 0.65), ovarian cancer (OR 0.53; 0.43, 0.66), small vessel stroke (OR 0.62; 0.47, 0.81), any ischemic stroke (OR 0.69; 0.61, 0.78), insomnia (OR 0.69; 0.64, 0.75), gout (OR 0.71; 0.60, 0.84), major depressive disorder (OR 0.78; 0.72, 0.85) and breast cancer (OR 0.85; 0.77, 0.94). Conversely, higher education level adjusted for intelligence was associated with an increased risk of obsessive-compulsive disorder (OR 2.24; 1.47, 3.41), bipolar disorder (OR 2.04; 1.64, 2.54), schizophrenia (OR 1.88; 1.49, 2.36), anorexia nervosa (OR 1.88; 1.53, 2.30) and anxiety (OR 1.84; 1.33, 2.56) (Figure 1). Results of sensitivity analyses were directionally similar but with wider CIs (Supplementary Figure 2). We detected possible pleiotropy in the analysis of obsessive-compulsive disorder, inflammatory bowel disease and rheumatoid arthritis (*p* for intercept in MR-Egger <0.05).

**Figure 1.**
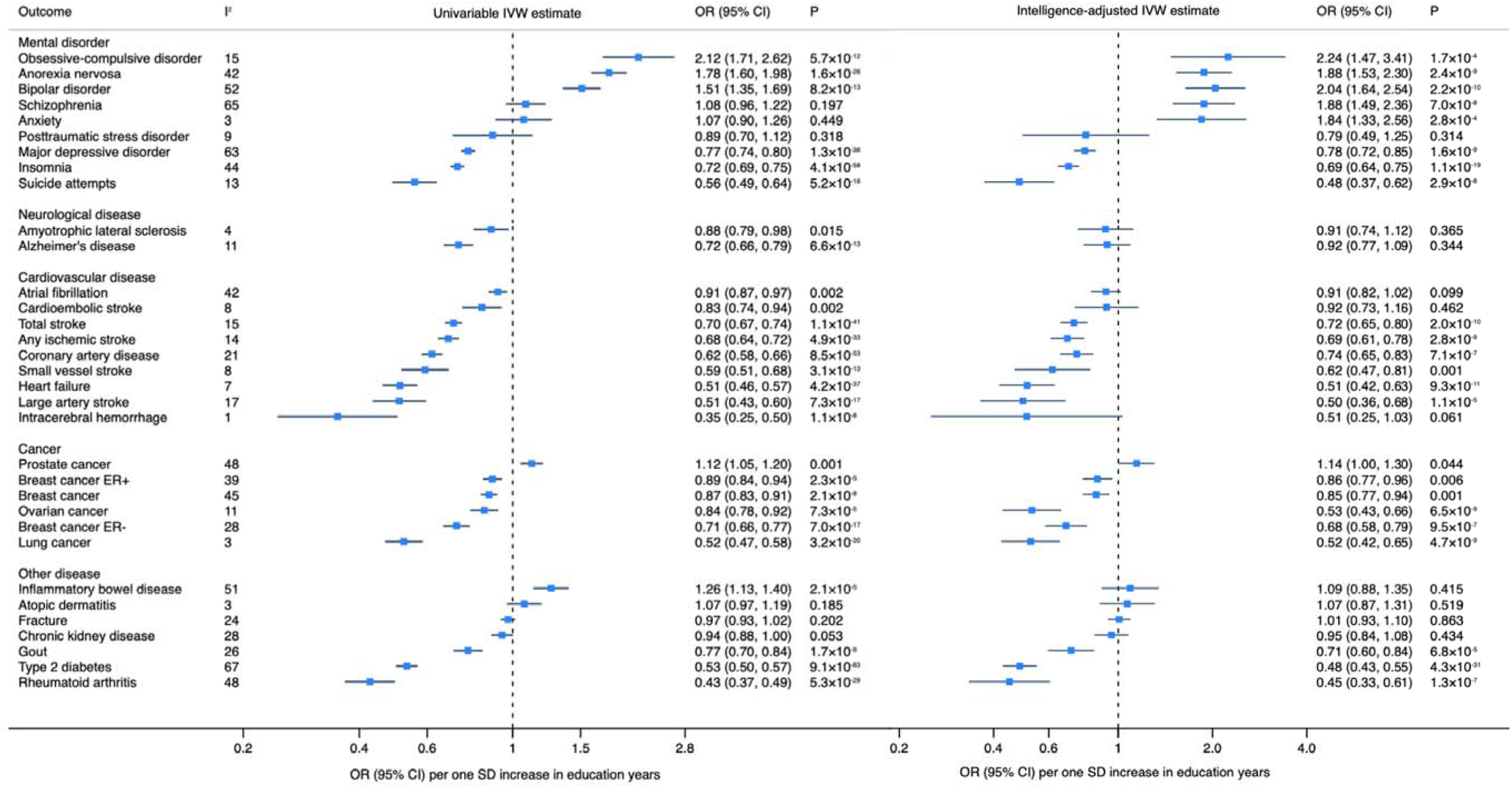
Associations of genetic predisposition to higher education level with health outcomes in MR analyses without and with adjustment for genetically predicted intelligence. CI indicates confidence interval; ER, estrogen receptor; IVW, inverse-variance weighted; OR, odds ratio; SD, standard deviation. I^2^ represents the degree of heterogeneity among included SNPs for education.

### Genetically predicted intelligence and diseases

The associations between intelligence and diseases are presented in Supplementary Figure 3 and 4. The effects of intelligence on mental and somatic diseases showed the same patterns with that of education level with the exception for schizophrenia (OR 0.47; 95% CI, 0.33, 0.67), bipolar disorder (OR 0.72; 95% CI, 0.52, 1.00), and Alzheimer’s disease (OR 0.77, 95% CI, 0.59, 1.01) after adjustment for education level (Supplementary Figure 3). Similar results were obtained from the weighted median method and pleiotropy was detected in the analysis of heart failure in the MR-Egger regression analysis (Supplementary Figure 4).

### Education, intelligence and risk factors

Genetically predicted higher education level was associated with later age of smoking initiation (β=0.31; 95% CI, 0.29, 0.33), higher bone mineral density (β=0.05; 95% CI, 0.03, 0.08), lower serum urate levels (β=-0.12; 95% CI, -0.15, -0.09), lower systolic (β=-0.13; 95% CI, -0.15, -0.11) and diastolic (β=-0.18; 95% CI, -0.20, -0.15) blood pressure, lower waist-to-hip ratio (β=-0.29; 95% CI, -0.31, -0.27), lower body mass index (β=-0.34; 95% CI, -0.37, 0.31) and fewer cigarettes smoked per day (β=-0.37; 95% CI, -0.42, -0.32) in the univariable model; the estimates were similar in the intelligence-adjusted model (Figure 2). Findings were consistent in sensitivity analyses and no pleiotropy was observed (Supplementary Figure 5). The same patterns of associations were observed for genetically predicted intelligence (Supplementary Figure 6). Nevertheless, after adjustment for education level, the magnitude of associations attenuated largely (Supplementary Figure 6).

**Figure 2.**
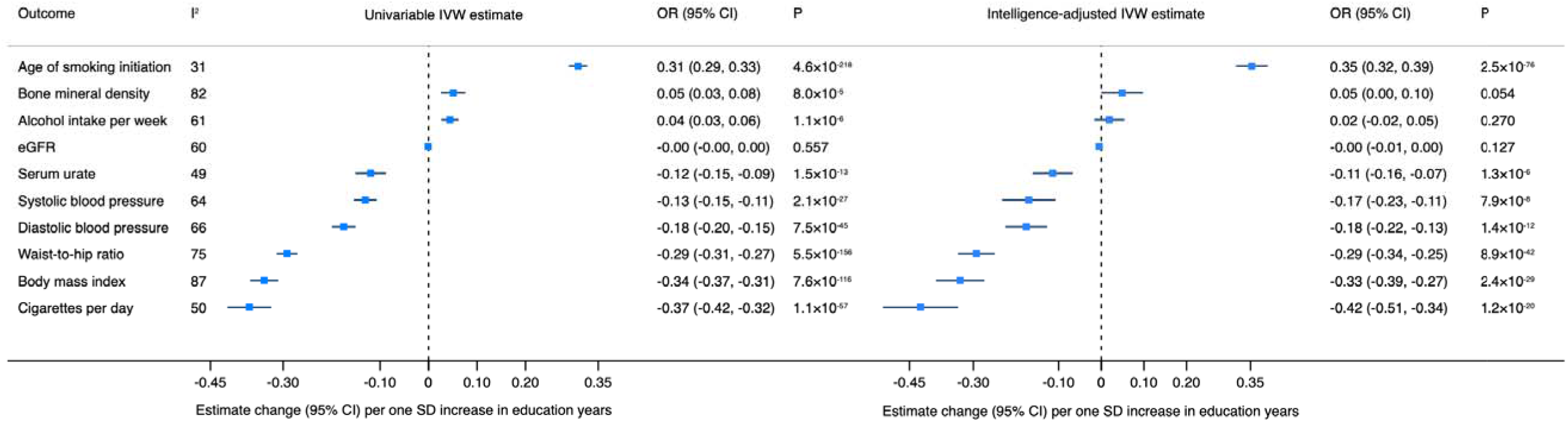
Associations genetic predisposition to higher education level and health-related risk factors in MR analyses without and with adjustment for genetically predicted intelligence. CI indicates confidence interval; eGFR, estimated glomerular filtration rate; IVW, inverse-variance weighted; SD, standard deviation. I^2^ represents the degree of heterogeneity among included SNPs for education.

### Comparison with observational studies

The present MR findings were generally similar in the direction and magnitude to the estimates based on meta-analyses of observational studies (Supplementary Table 3). However, there were discrepancies concerning the effects of education level on suicide attempts, breast cancer and prostate cancer.

### Mediation effects of body mass index and smoking

Table 2 shows the results of mediation analyses after adjusting for body mass index and smoking behaviour in the pathway from education to health outcomes. Although not apparent for all disease outcomes, body mass index and smoking partly mediated most associations between education and diseases (Table 2). A strong mediation effect of body mass index was observed in the associations of education with gout (68%), type 2 diabetes (57%) and heart failure (34%). With regard to smoking, a strong mediation effect was detected in the association of education with major depressive disorder (24%), type 2 diabetes (20%) and lung cancer (19%). After adjustment for both body mass index and smoking, the direct causal effect of education on outcomes was substantially attenuated for gout (64%), type 2 diabetes (63%), heart failure (36%), obsessive-compulsive disorder (32%), suicide attempts (31%), coronary artery disease (31%) and lung cancer (29%).

**Table 2.** Mediation analysis to disentangle the effects of body mass index and smoking in the pathway from education level to health outcomes

| Health outcome | Total effect of education |  |  | Effect after adjusting for BMI |  |  |  | Effect after adjusting for smoking |  |  |  | Effect after adjusting for both |  |  |  |
| --- | --- | --- | --- | --- | --- | --- | --- | --- | --- | --- | --- | --- | --- | --- | --- |
|  | OR<sup>a</sup> | 95% CI | P | OR<sup>b</sup> | 95% CI | P | %* | OR<sup>c</sup> | 95% CI | P | %* | OR<sup>d</sup> | 95% CI | P | %* |
| **Mental disorder** |  |  |  |  |  |  |  |  |  |  |  |  |  |  |  |
| Anorexia nervosa | 1.78 | (1.60, 1.98) | 1.56E-26 | 1.54 | (1.36, 1.75) | 1.70E-11 | 25 | 1.83 | (1.63, 2.05) | 3.00E-24 | 0 | 1.58 | (1.39, 1.80) | 4.90E-12 | 21 |
| Insomnia | 0.72 | (0.69, 0.75) | 4.15E-58 | 0.74 | (0.71, 0.78) | 2.80E-34 | 8 | 0.75 | (0.72, 0.79) | 1.10E-36 | 12 | 0.76 | (0.73, 0.80) | 1.80E-26 | 16 |
| Major depressive disorder | 0.77 | (0.74, 0.80) | 1.28E-36 | 0.80 | (0.76, 0.84) | 9.60E-20 | 15 | 0.82 | (0.78, 0.85) | 9.40E-19 | 24 | 0.83 | (0.79, 0.87) | 2.80E-13 | 29 |
| Obsessive-compulsive disorder | 2.12 | (1.71, 2.62) | 5.66E-12 | 1.70 | (1.31, 2.19) | 5.50E-05 | 29 | 1.99 | (1.57, 2.53) | 1.10E-08 | 8 | 1.67 | (1.28, 2.17) | 2.00E-04 | 32 |
| Suicide attempts | 0.56 | (0.49, 0.64) | 5.18E-18 | 0.64 | (0.54, 0.75) | 3.60E-08 | 23 | 0.61 | (0.52, 0.70) | 2.10E-11 | 15 | 0.67 | (0.57, 0.79) | 2.50E-06 | 31 |
| **Neurological disease** |  |  |  |  |  |  |  |  |  |  |  |  |  |  |  |
| Amyotrophic lateral sclerosis | 0.88 | (0.79, 0.98) | 1.50E-02 | 0.88 | (0.78, 1.00) | 4.60E-02 | 0 | 0.83 | (0.74, 0.93) | 1.00E-03 | 0 | 0.84 | (0.74, 0.95) | 8.00E-03 | 0 |
| Alzheimer's disease | 0.72 | (0.66, 0.79) | 6.60E-13 | 0.73 | (0.65, 0.81) | 4.17E-09 | 4 | 0.72 | (0.65, 0.79) | 3.00E-11 | 0 | 0.72 | (0.64, 0.80) | 5.01E-09 | 0 |
| **Cardiovascular disease** |  |  |  |  |  |  |  |  |  |  |  |  |  |  |  |
| Coronary artery disease | 0.62 | (0.58, 0.66) | 8.54E-53 | 0.70 | (0.65, 0.75) | 2.30E-21 | 25 | 0.66 | (0.61, 0.70) | 9.30E-34 | 13 | 0.72 | (0.67, 0.78) | 2.50E-17 | 31 |
| Heart failure | 0.51 | (0.46, 0.57) | 4.16E-37 | 0.64 | (0.56, 0.72) | 3.50E-13 | 34 | 0.54 | (0.48, 0.60) | 1.80E-26 | 8 | 0.65 | (0.57, 0.73) | 1.20E-11 | 36 |
| Total stroke | 0.70 | (0.67, 0.74) | 1.10E-41 | 0.75 | (0.71, 0.80) | 1.10E-19 | 19 | 0.71 | (0.67, 0.75) | 5.40E-33 | 4 | 0.76 | (0.71, 0.80) | 3.70E-18 | 23 |
| Any ischemic stroke | 0.68 | (0.64, 0.72) | 4.86E-33 | 0.70 | (0.65, 0.76) | 8.70E-20 | 8 | 0.68 | (0.64, 0.73) | 2.40E-27 | 0 | 0.70 | (0.65, 0.76) | 1.70E-18 | 8 |
| Large artery stroke | 0.51 | (0.43, 0.60) | 7.33E-17 | 0.55 | (0.46, 0.67) | 1.60E-09 | 11 | 0.53 | (0.44, 0.63) | 9.00E-13 | 6 | 0.56 | (0.46, 0.69) | 1.40E-08 | 14 |
| Small vessel stroke | 0.59 | (0.51, 0.68) | 3.07E-13 | 0.61 | (0.52, 0.73) | 1.40E-08 | 6 | 0.59 | (0.50, 0.69) | 2.00E-11 | 0 | 0.61 | (0.51, 0.72) | 2.60E-08 | 6 |
| **Cancer** |  |  |  |  |  |  |  |  |  |  |  |  |  |  |  |
| Breast cancer | 0.87 | (0.83, 0.91) | 2.12E-08 | 0.88 | (0.83, 0.94) | 4.00E-05 | 8 | 0.88 | (0.84, 0.93) | 8.80E-06 | 8 | 0.89 | (0.84, 0.95) | 3.00E-04 | 16 |
| Breast cancer ER+ | 0.89 | (0.84, 0.94) | 2.34E-05 | 0.90 | (0.84, 0.97) | 4.00E-03 | 10 | 0.90 | (0.85, 0.96) | 1.00E-03 | 10 | 0.91 | (0.85, 0.98) | 1.20E-02 | 19 |
| Breast cancer ER- | 0.71 | (0.66, 0.77) | 6.97E-17 | 0.70 | (0.64, 0.77) | 1.80E-13 | 0 | 0.72 | (0.66, 0.78) | 4.20E-17 | 4 | 0.71 | (0.64, 0.78) | 2.50E-12 | 0 |
| Lung cancer | 0.52 | (0.47, 0.58) | 3.20E-30 | 0.58 | (0.50, 0.66) | 1.30E-15 | 17 | 0.59 | (0.53, 0.67) | 6.30E-17 | 19 | 0.63 | (0.55, 0.72) | 2.90E-11 | 29 |
| Ovarian cancer | 0.84 | (0.78, 0.92) | 7.30E-05 | 0.90 | (0.81, 0.99) | 3.60E-02 | 40 | 0.82 | (0.75, 0.90) | 4.00E-05 | 0 | 0.88 | (0.79, 0.98) | 1.40E-02 | 27 |
| Prostate cancer | 1.12 | (1.05, 1.20) | 1.00E-03 | 1.06 | (0.98, 1.15) | 1.45E-01 | 49 | 1.07 | (1.00, 1.15) | 6.00E-02 | 40 | 1.04 | (0.96, 1.13) | 3.23E-01 | 65 |

| **Other diseases** |  |  |  |  |  |  |  |  |  |  |  |  |  |  |  |
| --- | --- | --- | --- | --- | --- | --- | --- | --- | --- | --- | --- | --- | --- | --- | --- |
| Gout | 0.77 | (0.70, 0.84) | 1.75E-09 | 0.92 | (0.84, 1.02) | 1.35E-01 | 68 | 0.76 | (0.69, 0.84) | 4.40E-08 | 0 | 0.91 | (0.81, 1.01) | 6.70E-02 | 64 |
| Rheumatoid arthritis | 0.43 | (0.37, 0.49) | 5.29E-29 | 0.41 | (0.34, 0.49) | 1.50E-21 | 0 | 0.44 | (0.37, 0.51) | 7.90E-23 | 3 | 0.42 | (0.35, 0.51) | 1.40E-19 | 0 |
| Type 2 diabetes | 0.53 | (0.50, 0.57) | 9.07E-83 | 0.76 | (0.71, 0.81) | 1.20E-12 | 57 | 0.60 | (0.56, 0.64) | 3.30E-48 | 20 | 0.79 | (0.74, 0.85) | 6.30E-11 | 63 |

BMI indicates body mass index; ER, estrogen receptor.
<sup>a</sup> total effect without any adjustment;
<sup>b</sup> adjusted for the effect of body mass index;
<sup>c</sup> adjusted for the effect of smoking (cigarettes per day);
<sup>d</sup> adjusted for the effects of both body mass index and smoking behaviors;
\*Percentage of the effect of education on the health outcome that is mediated by body mass index, smoking, or both (Formula: $\log(\text{OR}_{\text{total}}) - \log(\text{OR}_{\text{adjusted}}) / \log(\text{OR}_{\text{total}}) * 100$ ). We replaced the values with zero for those percentage below zero.

## Discussion

In the present MR study with up to 1.3 million individuals, genetic predisposition to higher education level was causally associated with the majority of major health outcomes and related risk factors. Specifically, genetic predisposition to higher education level, independent of intelligence, was associated with lower risk of major depressive disorder, insomnia, suicide attempts, stroke, heart failure, coronary artery disease, breast cancer, ovarian cancer, lung cancer, gout, type 2 diabetes, and rheumatoid arthritis. Conversely, higher education level was associated with higher risk of obsessive-compulsive disorder, anorexia nervosa, bipolar disorder, and prostate cancer. Genetically predicted higher intelligence, independent of education, was inversely related to bipolar disorder, schizophrenia and Alzheimer’s disease. Body mass index and smoking displayed strongest mediation effects observed for gout, type 2 diabetes, obsessive-compulsive disorder, suicide attempts and heart failure.

### Comparison with previous studies

Our findings are broadly in line with a vast body of observational studies showing a protective association of high educational level on major depressive disorder ^42^, amyotrophic lateral sclerosis ^45^, Alzheimer’s disease ^46^, coronary heart disease ^47^, heart failure ^48^, stroke ^49^, lung cancer ^52^, type 2 diabetes ^53^, chronic kidney disease ^54^, hypertension ^56^ and obesity ^55^. However, for suicide attempts, posttraumatic stress disorder, breast cancer and prostate cancer, our MR findings differ from observational findings. The discrepancies might be attributed by reverse causality in the observational studies, heterogeneity and small sample sizes in the meta-analyses. A substantial heterogeneity (I^2^=85%; *p*<0.001) was observed among included observational studies in the meta-analysis of breast cancer ^50^, and the sample size was small for prostate cancer ^51^. Some studies have proposed that the higher risk of prostate cancer among men with high education level was driven by higher prostate-specific antigen screening rate among educated men compared with men with low education level ^66^. With regard to the inverse associations of higher education level with breast and ovarian cancer, these associations may in part be mediated by reproductive or hormone-related factors, or other health behaviours such as healthier diet and physical activity. We are not aware of any previous MR studies on education or intelligence in relation to prostate, breast, or ovarian cancer, but a protective causal effect of higher education on lung cancer risk has been reported recently ^67^.

Previous MR studies based on much fewer SNPs (up to 162) as predictors of education level showed a protective effect of higher educational level on coronary artery disease ^68^ and Alzheimer’s disease ^69^. The present study based on substantially more SNPs as instrumental variables more precisely verified the findings in previous research. Notably, the effects of high education level in the previous studies might be influenced by high intelligence given the tight phenotype and genetic correlation between intelligence and education level. In the present study, we used multivariable MR analysis to assess the direct effect of education level that is not mediated via intelligence with coronary artery disease and Alzheimer’s disease. For Alzheimer’s disease, we found that higher intelligence rather than education level may be the protective factor, whereas higher education level was the protective factor for coronary artery disease. In a previous MR study of the direct effect of education and intelligence on certain health outcomes, including diabetes, hypertension, heart attack, total stroke, total cancer, and depression, no significant association with education or intelligence was observed despite significant or suggestive associations of genetically predicted education with potential risk factors (blood pressure, smoking, alcohol consumption, body mass index, vigorous physical activity, and television watching) ^59^. However, the genetic instrument used in that study comprised only 219 SNPs associated with education or intelligence and genetic associations with the outcomes were estimated in up to 138 670 UK Biobank participants, with relatively few outcomes. Thus, the power may have been insufficient to detect associations. Findings of other MR studies of education level in relation to health-related risk factors, such as obesity ^70^, cigarette smoking ^71^ and blood pressure ^72^, are consistent with our findings.

The inverse association between intelligence and schizophrenia was also observed in an observational study with 24 706 Swedish adults independent of overlapping genetic risks of two traits ^73^. However, the effects of high intelligence on bipolar disorder were conflicting across observational studies. A large-scale cohort study with over 20-year follow-up duration proposed a “reversed-J” shape association between intelligence and risk of bipolar disorder, which indicated that individuals with the lowest and highest intelligence had the greatest risk of bipolar disorder) ^74^. In a GWAS, it was revealed that bipolar disorder risk alleles were associated with better cognitive performance ^75^, which is opposite to our findings. Since education had a contradictory effect, which we observed in the present study, on bipolar disease to intelligence, the discrepancy among these studies might be caused by a mixed effect of education and intelligence at both aspects of phenotype and gene.

### Possible mechanisms

Based on results of the present MR study and previous observational studies, there are three major possible pathways linking education level to health outcomes: 1) modifiable risk factors largely mediates the educational effects on diseases ^72,76^; 2) there may be direct effects from education-related brain structures or function change via gene methylation, gene silencing etc. ^77-79^, especially for mental and neurological disorders; and 3) subjective wellbeing, happiness and meaning of life influenced by education level exerts effects on mental and somatic diseases directly or indirectly ^80-83^. Education, as measured in this study, can be defined as an institutionalized form of social resource, and more specifically a form of cultural capital drawing on the terminology of the French sociologist Pierre Bourdieu. Related forms of cultural capital emerge as objectivized resources – such as books, art or scientific tools – or incorporated resources, such as knowledge, attitudes and practices ^84,85^. Our study shows that education is a health relevant cultural capital whilst intelligence is not to the same degree related with health and risk of disease.

Observational studies have found that the associations between education level and diseases attenuated largely after adjustment for health-related risk factors. Compared with unadjusted model, the risk of cardiovascular diseases of low education attainment attenuated around 30-45% in statistical models adjusted for multiple risk factors, such as smoking, body mass index, hypertension and physical activity ^86,87^. However, measurement error and misclassification of mediators in observational studies often underestimates the mediation effects. The mediation effects were also proved in previous MR analysis ^72,76^. In the present study, genetically predicted education level was associated with a favourable risk factor profile: with improved smoking behaviours (postponed smoking initiation age and less cigarette per day) as well as lower adiposity (body mass index and waist-to-hip ratio), blood pressure and serum urate levels, which might mediate associations between education level and diseases. By conducting mediation analysis, we showed that body mass index and smoking behaviour partly or entirely mediated the pathway from education level to several health outcomes, in particular gout, type 2 diabetes, obsessive-compulsive disorder, suicide attempts, atrial fibrillation, heart failure, coronary artery disease and lung cancer.

Previous studies have found that low education level might influence the changes in biochemical response and risk-related brain function, such as inflammation ^77^, cardiometabolic traits ^78^ and amygdala reactivity ^79^, via gene methylation, thereby influencing disease risk. In addition, genetic studies have also revealed that improvement of subjective well-being ^80,81^, happiness ^80,81^, meaning of life ^82^, social interaction ^83^, possibly derived from high education level benefited human health directly and indirectly (e.g. influencing brain morphology, central nervous system and adrenal/pancreas tissues). There are other possible explanations, like followings: education level also could modify the risk of health outcomes through other diseases (comorbidity), the use of health care services, neighbourhood environment, occupations, income and marital status, which were amenable if education level was increased.

The results indicate that more than knowledge itself is affecting how people live their life, for instance through pathways regarding reduced smoking or alcohol habits among highly educated people. Therefore, we should consider further explanations, such as the relationship between high education on the one hand and the status and resources that follow it, on the other, which could by itself have a positive health effect on the individual. A further explanation assumes that it is the process itself that can be associated with increased wellbeing. That is, the process of taking part of and acquiring external knowledge rather than remaining with one’s own innate thinking or being kept oblivious. Should only a fraction of the disease burden be explained by this process of mental activity - given that education leads to a different kind of thinking, which is supported by the present study in that health is affected regardless of intelligence level - then increased knowledge through education may lead to longevity through mechanisms beyond health literacy pathways of late-onset diseases and beyond the influence of social and material factors.

### Strengths and limitations

The present study is the first study that comprehensively investigated the causal effects of education and intelligence on a very broad range of major disease outcomes and associated risk factors using genetic data from large-scale GWASs and genetic consortia. We used much more SNPs deriving from a larger GWAS with around 1.1 million individuals as instrumental variables for education level and the latest GWASs with largest sample size for outcomes compared to previous MR studies, thereby assuring adequate statistical power to detect weak associations. In addition, we disentangled the independent effect of education level from intelligence using a multivariable MR approach. Thus, it is a straightforward approach to estimate the possible health benefits from education promotion among general population. We used mediation analysis to reveal the roles of body mass index and smoking behaviour as mediators in the pathway from education level to health outcomes. Even though there were genetic data for certain outcomes from GWASs with trans-ancestry populations, the majority of included participants were individuals with European ancestry thereby diminishing population stratification bias. However, population confinement limited the transferability of the present findings to populations of non-European ancestries.

The major limitation of the present study is the possible unbalanced horizontal pleiotropy aroused from used genetic variants marking more generic biological pathways. It has been found that the lead SNPs related to education level and intelligence are significantly overexpressed in the central nervous system, such as hippocampus and cerebral cortex, but not in other organs ^40^. For cardiovascular disease, cancers and other physical diseases, we can minimize the possibility of pleiotropy from the global or systemic measures of fitness (such as mitochondrial function). It is more likely to conclude that the potential pleiotropy might exert a large to moderate effect via predominantly neurological pathways (for example, behaviours associated with obesity or smoking) for somatic diseases. In this scenario, the vertical pleiotropy would not bias the total causal effect by a higher educational level on disease development. With regard to mental and neurological disorders, although gene overwhelmingly expressed in the brain or central nervous system, studies found no, or at most a small, genetic correlation between lower education attainment and mental and neurological disorders by using bivariate genomic-relationship-matrix restricted maximum likelihood analysis ^1^. Thus, the associations between education level and mental or neurological diseases were not mainly because of measurable pleiotropic genetic effects, but because of education-related environmental factors. In addition, from a statistical perspective, we detected almost no pleiotropy in the results of MR-Egger regression and the estimates were consistent through sensitivity analyses, which indicated a negligible distortion by pleiotropy. Intergenerational effects from parents for certain disease, such as coronary artery diseases and type 2 diabetes, could not be assessed by using the data in the present MR study.

In summary, the present MR study strengthened the evidence of protective role of high education level on the majority of mental disorders and somatic diseases independent of intelligence. Body mass index and smoking partly mediated several of the associations between education level and health outcomes. These findings strongly suggest increasing education level for overall health benefits.

## Data Availability

The datasets analysed in this study are publicly available summary statistics.

## Acknowledgments

Summary-level data for education level, intelligence and outcomes were obtained from the UK Biobank and genetic consortia, including Psychiatric Genomics Consortium (bipolar disorder, schizophrenia, obsessive-compulsive disorder, post-traumatic stress disorder, anxiety and anorexia nervosa), The International Genomics of Alzheimer’s Project (IGAP), Project MinE, Center for Neurogenomics and Cognitive Research (CNCR), iPSYCH, CARDIoGRAMplusC4D Consortium, MEGASTROKE Consortium, Atrial Fibrillation Consortium (AFGen), International Stroke Genetics Conosrtium (ISGC), Breast Cancer Association Consortium (BCAC), The Prostate Cancer Association Group to Investigate Cancer Associated Alterations in the Genome (PRACTICAL) consortium, The International Lung Cancer Consortium (ILCCO), The Ovarian Cancer Association Consortium (OCAC), The DIAbetes Genetics Replication And Meta-analysis (DIAGRAM) consortium, The Global Urate Genetics Consortium (GUGC), Genetics and Allied research in Rheumatic diseases Networking (GARNET) consortium, UK Inflammatory Bowel Disease Genetics (IBD) Consortium, CKDGen Consortium, the EArly Genetics and Lifecourse Epidemiology (EAGLE) Consortium, GEnetic Factors for OSteoporosis (GEFOS) Consortium, GIANT consortium, Neale lab, Consortium of Alcohol and Nicotine use (GSCAN). The authors thank all investigators for sharing these data. The list of investigators of MEGASTROKE is available at http://megastroke.org/authors.html. Funding of the MEGASTROKE project are specified at megastroke.org/acknowledgements.html.

## Author contributions

SY drafted the manuscript and conducted the statistical analyses. SY and YX conducted the systematic review. All authors contributed to the interpretation of the results and critical revision of the manuscript for important intellectual content and approved the final version of the manuscript.

## Ethical approval

This MR study was approved by the Swedish Ethical Review Authority.

## Informed consent

All participants included in the genome-wide association studies gave informed consent.

## Data sharing

The datasets analyzed in this study are publicly available summary statistics.

